# A haplotype-based approach for myotonic dystrophy type 1 identification

**DOI:** 10.64898/2026.07.09.26357389

**Authors:** Claudia Moreau, Gilles-Philippe Morin, Joanie Bouchard, Jean Mathieu, Elise Duchesne, Cynthia Gagnon, Simon L. Girard

## Abstract

**Background:** Myotonic dystrophy type 1 (DM1) is caused by a CTG repeat expansion in the *DMPK* gene and represents the most common adult-onset myopathy. Current molecular diagnostics rely on labor-intensive assays that limit accessibility and scalability. Haplotype-based approaches offer a promising alternative for detecting pathogenic expansions indirectly.

**Methods:** We performed genome-wide genotyping in 226 genetically confirmed DM1 patients from the Saguenay–Lac-Saint-Jean founder population and reconstructed haplotypes surrounding the *DMPK* pathogenic repeat expansion. Based on these haplotypes, we performed a phylogenetic analysis that was further integrated with genealogical reconstruction from the BALSAC database to investigate the origin and transmission of DM1 haplotypes. To evaluate epidemiological utility, we implemented gene dropping simulations within the SLSJ extended genealogies (>80,000 starting individuals) to estimate DM1 incidence at birth.

**Results:** A DM1-associated haplotype was identified in all patients (226/226), consistent with a single major ancestral origin in the SLSJ population. This complete concordance supports the robustness of haplotype-based approaches to infer carrier status without direct repeat sizing. Integrating phylogenetic analysis and genealogical data identified a single couple as the most likely entry point of DM1 in Quebec. Simulation-based estimates of incidence at birth exceeded observed prevalence, suggesting underdiagnosis in the region. Marked geographic heterogeneity in the SLSJ is also observed.

**Conclusions:** Our results demonstrate that haplotype-based approaches can provide a reliable, cost-effective alternative to conventional pathogenic DM1 repeat carriers identification and familial screening strategies.

## Background

Myotonic dystrophy type 1 (DM1) (MIM: 160900) is the most common form of muscular dystrophy in adults and is characterized by progressive myopathy, myotonia, and multiorgan involvement (1,2). It is an autosomal dominant disorder characterized by a wide range of ages at symptom onset. Due to the marked variability in clinical presentation, individuals with DM1 and their caregivers often face substantial financial, logistical, and accessibility barriers, contributing to a prolonged diagnostic odyssey (3). Life expectancy is greatly reduced in DM1 patients, particularly in those with early onset and more severe phenotype (4).

DM1 is caused by an expansion of at least 50 CTG repeats in a non-coding region of the *DMPK* gene, sometimes reaching several thousand repeats (5). The molecular diagnosis is typically performed using southern blot analysis, which is labor intensive and costly (CAD 425 per sample in Quebec). In addition to barriers to genetic testing, studies have reported substantial diagnostic delays in DM1, driven by its multisystem and heterogeneous presentation and limited disease recognition outside neuromuscular specialties. In consequence, DM1 genetic diagnosis is not uniformly accessible to affected families (5). Improving diagnostic strategies for DM1 is becoming increasingly critical for the patients’ management (6) in the context of ongoing clinical trials and the potential introduction of disease modifying therapies. Moreover, recent newborn screening data suggest that DM1 may be up to five times more common than previously reported, reinforcing the need for rapid screening of at-risk family members (7).

Improving the accessibility of DM1 identification and family screening will likely require the adoption of alternative strategies, such as haplotype-based approaches. A haplotype is defined as a set of genetic variants that are physically close and therefore tend to be inherited together. Rather than investigating individual variants in isolation, haplotype-based approaches leverage these correlated blocks of variation that can be shared among individuals. Haplotype based strategies have been successfully applied in prenatal diagnosis (8–10), carrier screening (11), and in clinical settings where direct mutation detection is technically challenging or costly (12,13). Interestingly, it has been shown that DM1 patients of diverse ancestry share a common haplotypic background (14,15). It is the case in the Saguenay–Lac-Saint-Jean (SLSJ) region of Quebec which has one of the highest reported prevalence of DM1 worldwide (158 per 100,000) (16) due to a strong founder effect. This well-characterized founder population also benefits from uniquely rich genealogical, genomic, and clinical data resources, making it an ideal model for assessing the feasibility and potential impact of haplotype-based testing for DM1.0

In this study, we characterized the haplotypes of 226 DM1 patients from the SLSJ region and evaluated whether haplotype analysis could be used for DM1 identification in family screening. We also aimed to trace the haplotype in the population through deep genealogies and to assess the expected incidence at birth in the whole SLSJ population.

## Data and Methods

### Data

For more than fifty years, the Neuromuscular Clinic of the Centre intégré universitaire de santé et de services sociaux (CIUSSS) of SLSJ has documented over 500 DM1 patients diagnosed through clinical evaluation and molecular testing (16). The SLSJ population is the subject of an ongoing longitudinal study since 2002 to document the severity and progression of physical, cognitive, pulmonary, cardiac impairments along with limitations of activities and restriction of social participation and identify their explanatory and predictor factors (17–21). The CTG repeat length was measured for more than 200 of these patients.

Multigenerational familial data of 1,428 DM1 patients and unaffected relatives were used to reconstruct genealogies with the BALSAC population registry (22), which was built on civil records since the beginning of the Quebec European settlement up to 1965. These were merged with genealogical data of 88,848 individuals who married in the SLSJ region between 1931-60 (23). The BALSAC database also includes all children born to couples married in the SLSJ since the beginning of the European settlement.

### Genotyping

We performed whole-genome genotyping on the Global Screening Array-24 Kit from blood for 226 DM1 patients from 58 families. Data was cleaned using PLINK software v1.9 (24), ensuring individuals with at least 95% genotypes among all SNPs were retained. At the SNP level, we retained SNPs with at least 95% genotypes among all individuals, removing A to T and G to C genotypes, located on the autosomes and in Hardy-Weinberg equilibrium p > 10^-6^. In total, we retained 476,009 SNPs genome-wide for 226 patients.

### Identity-by-descent and haplotyping

We inferred identity-by-descent (IBD) segments on phased genotypes using Refined IBD (25) version 17Jan20 and Beagle version 5.1. We retained only IBD segments of 2 Mb or longer and with a LOD score greater than 3. We then examined the proportion of individuals’ pairs sharing IBD along the genome for all patients. This proportion is around 0.02 in SLSJ individuals who are not closely related (26). We detected a peak of sharing around the repeat on the *DMPK* gene as expected (Supplementary Fig 1). Based on this level of sharing, we defined two haplotype boundary sets spanning 2 Mb where 35% of pairs shared an IBD segment, and 17 Mb where 10% of pairs shared IBD using the R package geneHapR (27). We determined linkage disequilibrium (LD) blocks within haplotype boundaries using Haploview v4.2 (28) for visualization purposes. For each patient, we defined the DM1 haplotype as the one shared with other patients. To validate the specificity of DM1 associated haplotypes, we compared them with the other haplotype of each patient (Supplementary Fig 2).

### Phylogeny of haplotypes

To reconstruct the history of DM1 haplotypes in the SLSJ population, we extended the haplotypes to 17 Mb. At this scale, most haplotypes resolve into family-specific haplotypes, although some recombination events remain. Given the dominant inheritance of DM1, segregation patterns within families are readily apparent, facilitating the grouping of similar haplotypes and the identification of recombination events. To limit the complexity associated with single recombination events in downstream phylogenetic analyses, we restricted our study to the 10 most frequent haplotypes, encompassing 45 individuals from 11 families. For each family, the closest common ancestor (CCA) was determined from genealogical data within a maximum of four generations. These familial CCA were used as the starting points for phylogenetic reconstruction, and each was assigned the most common haplotype observed among their descendants, eliminating unique recombination events. Phylogenetic inference was then performed using IQ-TREE v2.3.6 (29) on the variable genotype positions spanning 17 Mb around the CTG repeat expansion on chromosome 19. We extracted the three valid trees with approximately unbiased (AU) test ≥ 0.05 for subsequent analyses.

### Alignment with the genealogy and inferred carrier frequency in SLSJ

These valid trees were aligned to the genealogy using the coaling tool (30) to find the ancestors who most probably introduced and propagated the DM1 repeat expansion (or susceptibility to expansion) in the SLSJ population.

### Incidence at birth inferred from simulations

Using clinical data from all patients and their relatives, and the genealogical path to the ancestor, we performed 1,000 gene dropping simulations, propagating DM1 alleles one generation forward with a 50% transmission probability to all children in the descending path. This was done using the ascending genealogy of all individuals married in the SLSJ between 1931-60 merged with DM1 patients. For all individuals who married in the SLSJ region, complete information on their offspring was also available. Expected DM1 incidence at birth was calculated as the proportion of children inheriting the simulated allele among all children born to women married in the SLSJ between 1931 and 1945. We limited the analysis to marriages occurring up to 1945 to avoid data truncation, as records end in 1965, which would lead to under-ascertainment of children born to parents married after 1945.

## Results

### Haplotyping

A DM1-associated haplotype was identified in 100% (226/226) of patients with genotype data (Fig. 1).

**Figure 1:**
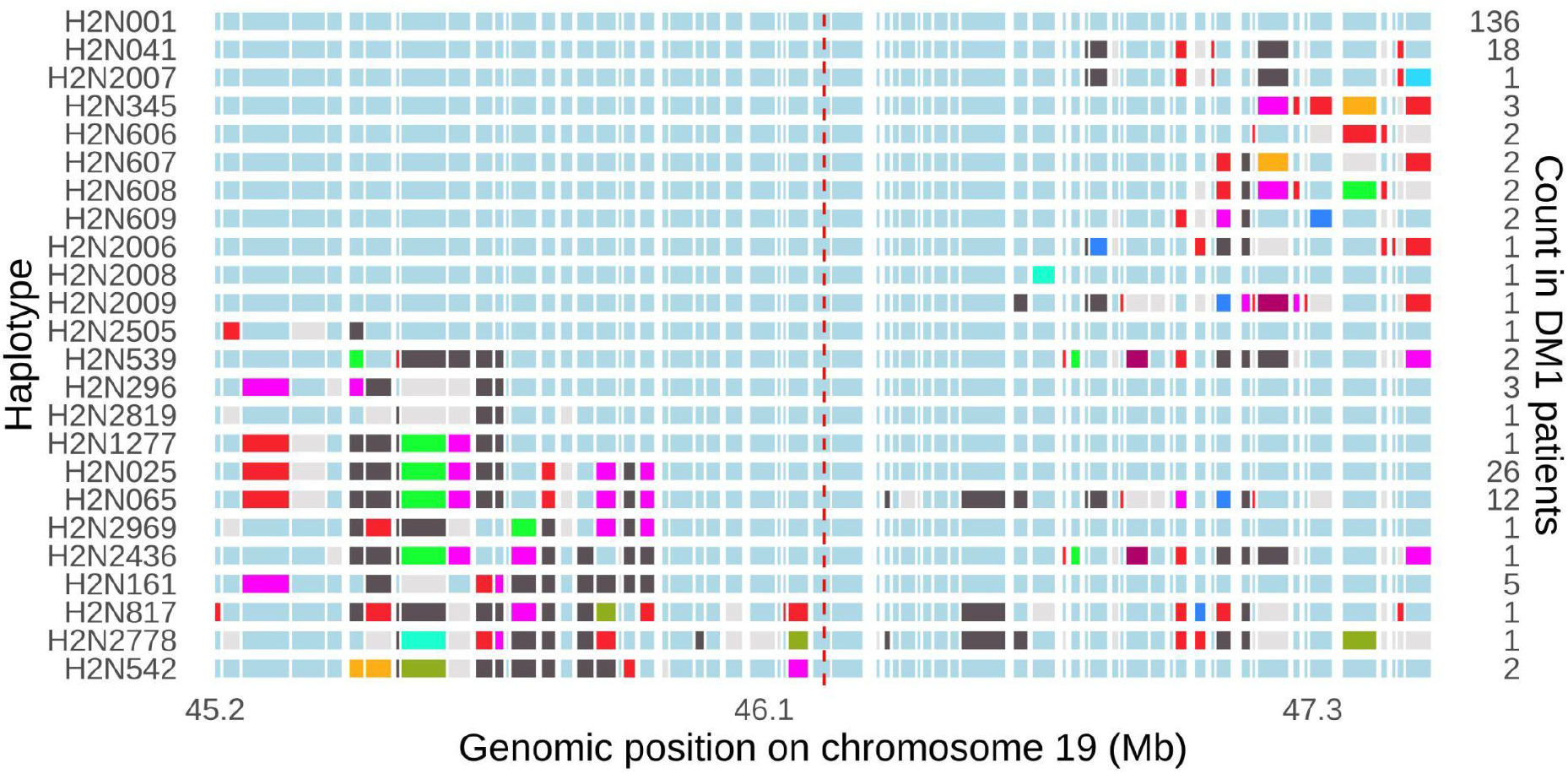
DM1-associated haplotype reconstruction. 2 Mb haplotypes are shown on the left y-axis and chromosomal positions on the x-axis. The count of each haplotype in the DM1 cohort is shown on the right y-axis. Instead of individual variant positions, LD blocks are displayed. A uniform color was applied across all haplotypes for identical LD blocks. The reference haplotype was selected as the most frequent among patients. The dashed red line shows the DM1 repeat expansion position on the *DMPK* gene.

To assess the specificity of the DM1 haplotype, we examined the second haplotype carried by each patient for evidence of shared structure with the DM1 haplotypes (Supplementary Fig. 2). No clear pattern surrounding the repeat was observed among these non-DM1 haplotypes.

### Phylogeny of haplotypes

After extending the DM1 haplotypes to 17 Mb, we identified the closest common ancestors (CCA) among families sharing identical or similar haplotypes. Only the major haplotype for each family was used in the phylogenetic analyses (see Methods), revealing three distinct possible topologies (Fig. 2).

**Figure 2:**
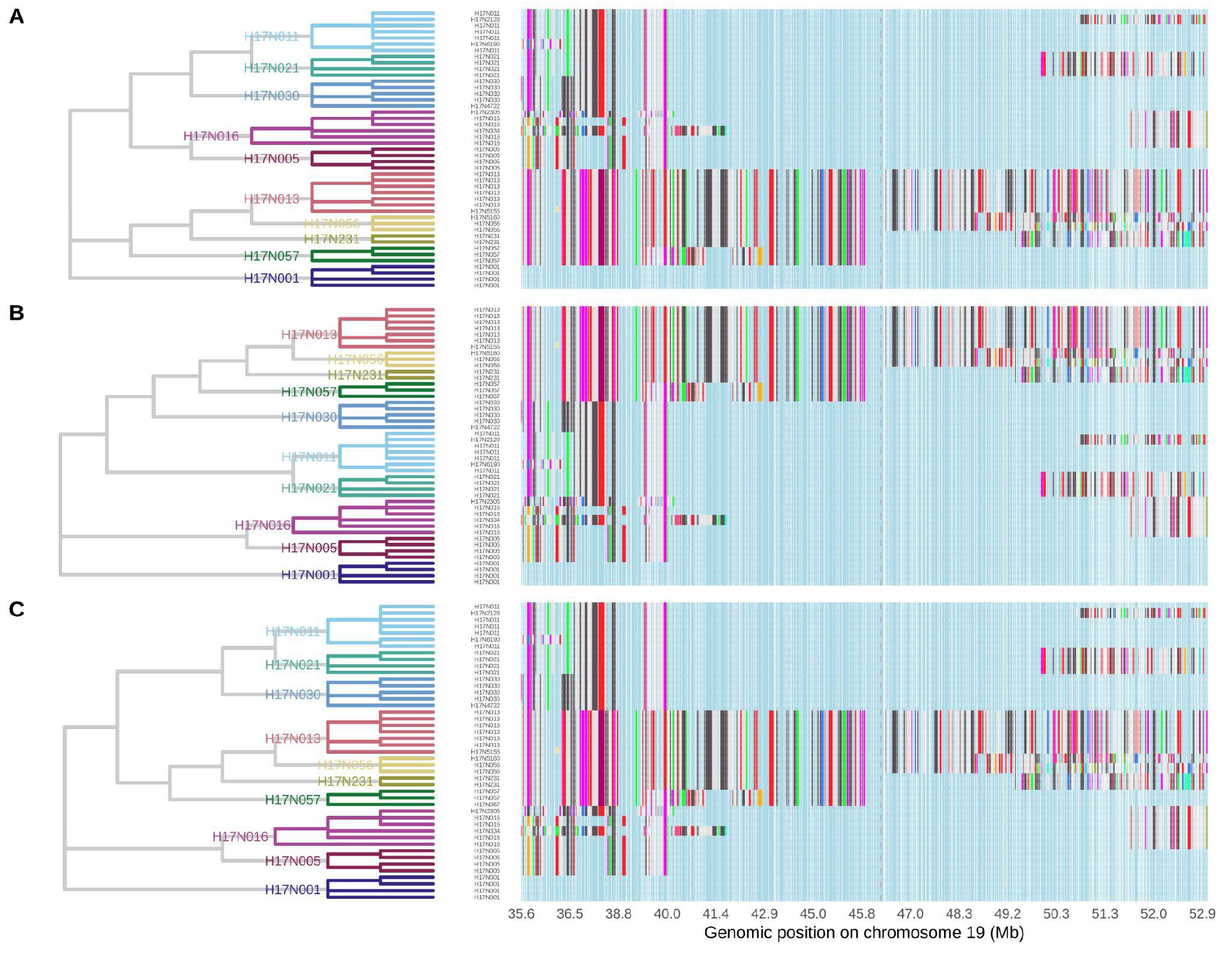
Alternative phylogenetic configurations of major DM1 haplotypes. Results are shown for the three statistically supported tree topologies (A to C). On the left, the possible phylogenetic reconstructions inferred using IQ-TREE are depicted by gray branches, extending from the familial closest common ancestors (CCAs) to the root. Colored branches represent the genealogical paths from DM1 patients to their respective CCA, with each color corresponding to a distinct family. Each CCA was assigned the family’s major haplotype. On the right, 17Mb DM1 haplotypes are shown on the y-axis and chromosomal positions on the x-axis. Instead of individual variant positions, LD blocks are displayed. A uniform color was applied across all haplotypes for identical LD blocks. The reference haplotype was selected as the most frequent among DM1 patients. The dashed red line shows the DM1 repeat expansion position on the *DMPK* gene.

### Alignment with the genealogy

We then aligned the three alternative phylogenetic trees with the genealogy of DM1 patients, starting from the CCAs. This approach identified three ancestral couples as potential introducers of the DM1 repeat expansion (or susceptibility to expansion) in Quebec. We selected the most probable ancestral couple (Supplementary Table 1) and reconstructed a possible DM1 haplotype transmission pathway to most patients based on haplotype structure and family data (Supplementary Fig 3). Genealogical validation confirmed that almost all patients excluded from this alignment nonetheless descended from the identified ancestral couple. The only individual who does not descend from this couple instead traces descent through the husband’s parents.

### Incidence at birth

Integrating the genealogies of all individuals married in the SLSJ between 1931 and 1960 (23) with those of DM1 patients, we performed gene dropping simulations to assess the DM1 incidence at birth for children of women married in the SLSJ between 1931-45. The mean incidence at birth for 1,000 simulations for this time period was 169 per 100,000 births. Notably, a marked geographic heterogeneity was observed across SLSJ, with the North of the Saguenay River showing the highest expected incidence at birth (Fig 3).

**Figure 3:**
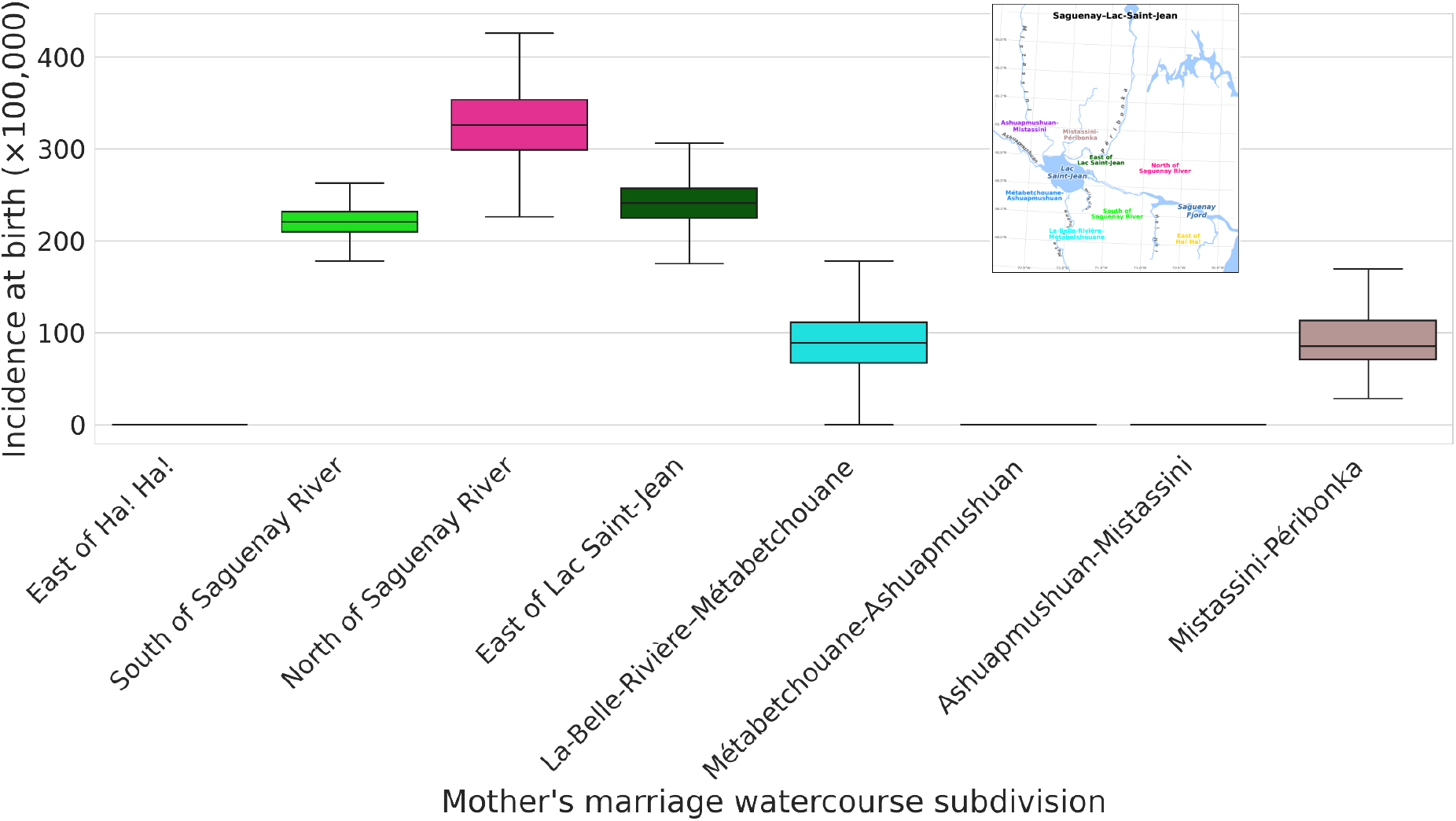
Simulated DM1 incidence at birth (1931–1945) by SLSJ subdivision. The incidence at birth was inferred from gene dropping simulations for women married between 1931-45 in each SLSJ subdivision. The subdivisions were taken from (23).

## Discussion

In this study, we identified a DM1 haplotype in 100% of the 226 genotyped patients from the SLSJ population. Our findings suggest that a haplotype-based approach could provide a simple, robust, and cost-effective alternative for DM1 identification and affected family screening.

Haplotype-based testing for DM1 may have utility beyond the SLSJ region and could be scalable to DM1 patients in other parts of the world. A previous study has shown that DM1 patients of diverse ancestry share a common haplotype background, suggesting that the original mutation event occurred in a population ancestral to all non-African populations (14). These observations suggest that haplotype-based DM1 testing could represent a broadly applicable strategy, extending beyond founder populations to diverse cohorts worldwide for DM1 identification and prevention. Moreover, haplotype-based approaches may be broadly applicable to repeat expansion disorders, as haplotypes have been identified across multiple repeat diseases (13,31–33).

We estimated the population-level DM1 incidence at birth to be 169 per 100,000 births. Previously reported DM1 prevalence estimates in the SLSJ region range from 142 to 178 per 100,000 individuals for the period 1985-2010 (16). Prevalence is generally expected to exceed incidence at birth, as it reflects the cumulative burden of affected individuals over time. However, prevalence could not be directly estimated in our study due to the lack of mortality data in the BALSAC database. Although our incidence estimate encompasses all individuals born to women married between 1931 and 1945, not all DM1 families were captured in our data. Nevertheless, some of those lost families were likely indirectly represented via shared affected ancestors within the genealogical lineages connecting patients to the ancestral couple presumed to have introduced the variant. As a result, the DM1 birth incidence estimate is likely underestimated, suggesting that its prevalence in the SLSJ population may also be higher than currently reported, as also observed in other populations (7).

We observed a geographic heterogeneity in the simulated incidence of DM1 at birth across SLSJ subdivisions (23), suggesting a spatial distribution which in part reflects that of identified DM1 patients (34). Consequently, individuals in some subdivisions may be at increased genetic risk of DM1 despite having no known family history of the disease. The instability of the DM1 CTG repeat expansion across generations results in anticipation, characterized by progressively earlier onset and greater disease severity (35,36) which may allow mildly affected or asymptomatic families to remain undetected. Consistent with this observation, new DM1 families continue to be identified at the CIUSSS du SLSJ, suggesting that a proportion of affected lineages remain unrecognized.

This study has several limitations. First, we lack systematic assessment of CTG repeat length in population controls, which would be informative for refining the specificity of DM1 haplotypes in the general population. Second, our approach to ancestor identification relies on accurate reconstruction of haplotype phylogenies, which is inherently challenging when including familial data; however, the availability of detailed genealogical information in this population partially mitigates this limitation. In addition, there is a generational gap between the genealogical data used for simulations of incidence at birth and the period over which prevalence has been observed. However, demographic expansion and the founder effect in SLSJ largely stabilized by the 1960s (37), suggesting that major differences in incidence or prevalence across this interval due to population growth are unlikely.

## Conclusions

In conclusion, haplotype-based DM1 identification of pathological expansion carriers provides a complementary framework to conventional molecular testing by enabling the early identification of at-risk individuals in families and lineages, including those who are currently asymptomatic or unaware of their carrier status. By facilitating the identification of affected individuals and at-risk relatives, these strategies could accelerate clinical trial recruitment and generate more reliable population-wide prevalence estimates in research settings. As DM1 therapies transition toward clinical implementation, integrating robust haplotype-based tools into clinical practice may prove essential for guiding eligibility, and ensuring equitable access to emerging precision medicine interventions.

## List of abbreviations

CCA: closest common ancestor
CIUSSS: Centre intégré universitaire de santé et de services sociaux
DM1: myotonic dystrophy type 1
IBD: identity-by-descent
LD: linkage disequilibrium
SLSJ: Saguenay–Lac-Saint-Jean

## Declarations

### Ethics approval and consent to participate

This project is approved by the Centre intégré universitaire de santé et de services sociaux (CIUSSS) du Saguenay–Lac-Saint-Jean and Université du Québec à Chicoutimi ethics boards.

### Consent for publication

Not applicable.

### Availability of data and materials

The datasets generated and analysed during the current study are not publicly available due [to the informed consent given by study participants but are available from the corresponding author on reasonable request.

Access to the BALSAC genealogical data is controlled in accordance with the Policy on Access to BALSAC Data for Research Purposes to protect participant confidentiality and adhere to ethical guidelines. The procedure for requesting access to BALSAC data is described at https://www.balsac.ca/en/data-infrastructure/. Access requests must be submitted to the Researchers Service of the BALSAC Project and must include a completed BALSAC Database Access Request Form along with all documents necessary for evaluation, such as a certificate of ethics approval. Applications are reviewed to ensure compliance with the scientific method, feasibility, and BALSAC access policies. Applicants are informed of the decision in writing. Questions regarding data access can be addressed to. If access is granted, data may be used only within the scope of the approved research project and must be destroyed once the authorized access period expires. The BALSAC data are not publicly available due to ethical restrictions on the publication of genealogical information dating back less than 100 years.

### Authors’ contributions

All authors approved the final version of the manuscript. CM played an important role in interpreting the results. GPM contributed in analyzing the genealogical data, CM and SLG conceived and designed the study and drafted the manuscript. CM and GPM performed the analyses. JB, ED and CG computerized the pedigree information for genealogical reconstruction. GPM, JB, JM, ED, CG revised the manuscript. JM was the principal investigator of the Natural history study and CG was a doctoral student at the time.

## Acknowledgements

We are extremely grateful to all participants in this research. This work was made possible by the Digital Research Alliance of Canada which provided access to storage and computing resources.

We thank Josée Villeneuve for her support in the clinical file review and genetic diagnosis of all patients. We also thanks Stéphanie Cloutier and Mélissa Verreault for

