## Supplementary material for "A haplotype-based approach for myotonic dystrophy type 1 identification"

#### Supplementary material
#### Figures

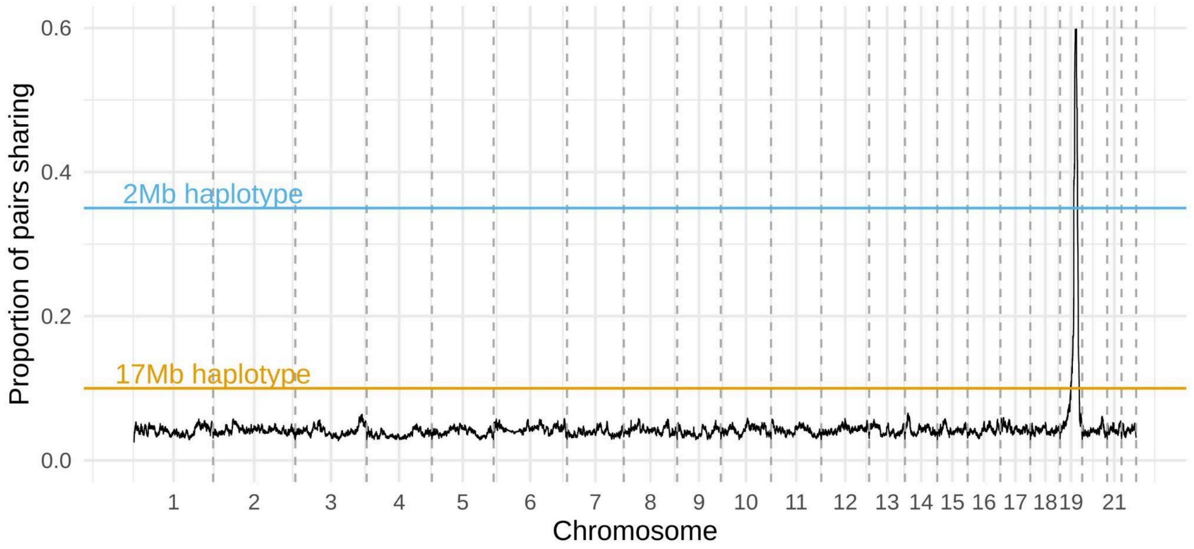


Supplementary Figure 1: **Proportion of patients' pairs sharing IBD at each genomic position.** A peak of sharing is observed around the *DMPK* gene on chromosome 19. The boundaries of a 2Mb and a 17Mb haplotypes were determined at genomic positions where 35% and 10% of patients shared IBD, respectively.

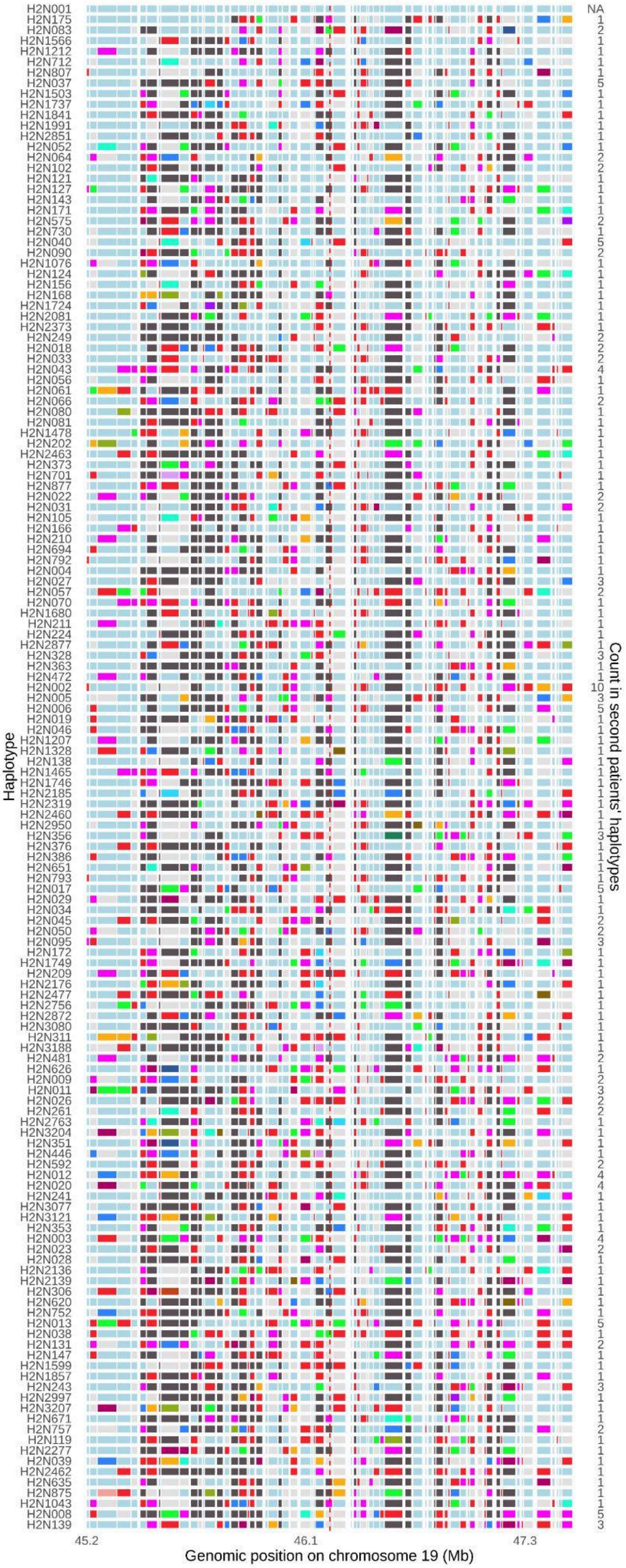

Haplotype
Count in second patients' haplotypes
45.2
46.1
47.3
Genomic position on chromosome 19 (Mb)

Supplementary Figure 2: **Non-DM1-associated haplotype reconstruction**. 2 Mb haplotypes are shown on the left y-axis and chromosomal positions on the x-axis. Counts of non-DM1-associated haplotypes are shown on the right y-axis. Instead of individual variant positions, LD blocks are displayed. A uniform color was applied across all haplotypes for identical LD blocks. The reference haplotype was selected as the most frequent associated with DM1 for comparison purposes. The dashed red line shows the DM1 CTG repeat position on the *DMPK* gene.

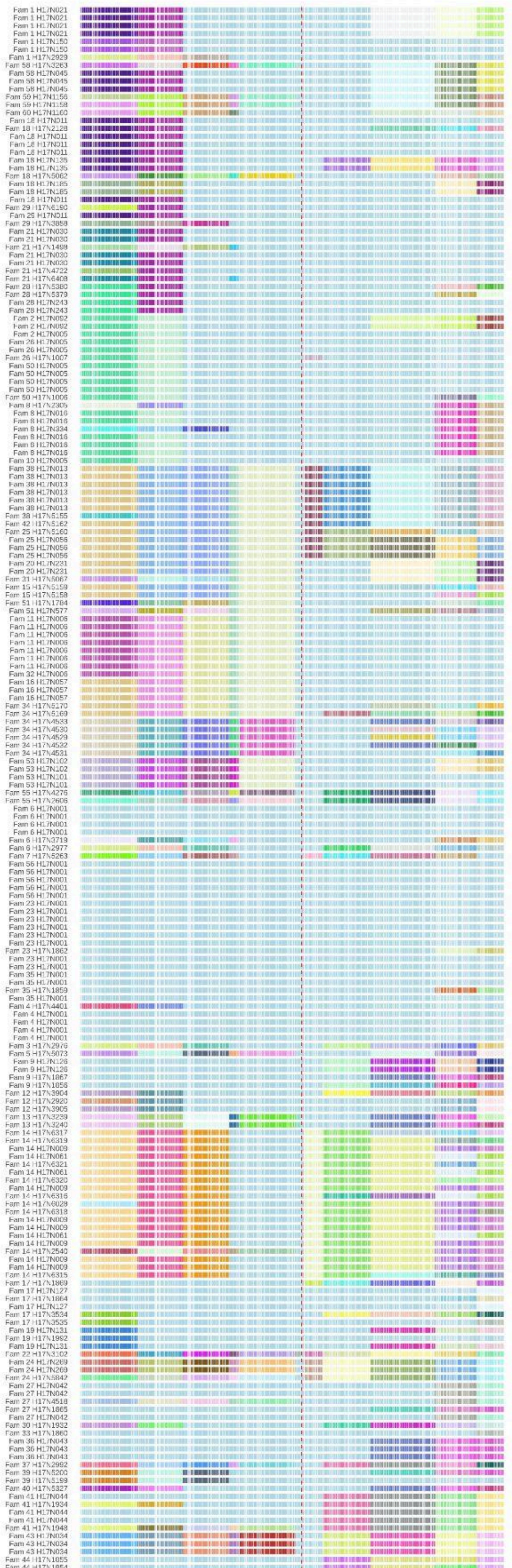
35.6
36.5
38.8
40.0
41.4
42.9
45.0
45.8
47.0
48.3
49.2
50.3
51.3
52.0
52.9
Genomic position on chromosome 19 (Mb)

Supplementary Figure 3: **DM1-associated haplotype reconstruction clustered by families and haplotype structure**. 17 Mb haplotypes are shown on the y-axis and chromosomal positions on the x-axis. Family identifiers were modified to protect confidentiality. Instead of individual variant positions, LD blocks are displayed. A uniform color was applied across all haplotypes for identical LD blocks' adjacent sequences. The reference haplotype was selected as the most frequent associated with DM1 (H17N001) for comparison purposes. The dashed red line shows the DM1 repeat expansion position on the *DMPK* gene.

### Table

Supplementary Table 1: **Probability of each ancestors' couple within each tree.**

| Ancestors' couple | Tree A | Tree B | Tree C |
|---|---|---|---|
| **Couple 1** | **0.988** | 0 | 0 |
| Couple 2 | 0.010 | 0.84 | 0 |
| Couple 3 | 0.002 | 0.16 | 1 |

Probabilities were calculated independently for each tree. Trees refer to figure 2 in the main text. The husband in couple 1 is a child of couple 2. Couple 1 was taken as the most probable couple having introduced the DM1 haplotype in Quebec.
